# Do people reduce compliance with COVID-19 guidelines following vaccination? A longitudinal analysis of matched UK adults

**DOI:** 10.1101/2021.04.13.21255328

**Authors:** Liam Wright, Andrew Steptoe, Hei Wan Mak, Daisy Fancourt

## Abstract

**Introduction:** COVID-19 vaccines do not confer immediate immunity and vaccinated individuals may still be at risk of transmitting the virus. Governments have not exempted vaccinated individuals from behavioural measures to reduce the spread of COVID-19, such as practicing social distancing. However, vaccinated individuals may have reduced compliance with these measures, given lower perceived risks.

**Methods:** We used monthly panel data from October 2020 – March 2021 in the UK COVID-19 Social Study to assess changes in compliance following vaccination. Compliance was measured with two items on compliance with guidelines in general and compliance with social distancing. We used matching to create comparable groups of individuals by month of vaccination (January, February, or not vaccinated by February) and fixed effects regression to estimate changes in compliance over the study period.

**Results:** Compliance increased between October 2020 – March 2021, regardless of vaccination status or month of vaccination. There was no clear evidence that vaccinated individuals decreased compliance relative to those who were not yet vaccinated.

**Conclusion:** There was little evidence that sample members vaccinated in January or February reduced compliance after receiving vaccination for COVID-19. Continued monitoring is required as younger individuals receive the vaccine, lockdown restrictions are lifted and individuals receive second doses of the vaccine.

## INTRODUCTION

Governments have begun mass vaccination programmes for COVID-19, but it will be several months before herd immunity is achieved. The available vaccines do not confer immediate immunity and are not 100% effective [1]. Vaccinated individuals may still be at risk of catching and transmitting the virus, including variants they have not been inoculated against [2]. Given this, the UK government has not exempted vaccinated individuals from behavioural measures to reduce the spread of COVID-19, such as the wearing of masks, practicing social distancing, and reducing household mixing.

International data show that, though compliance levels are high overall, not all individuals comply with recommended or mandated behavioural measures [3]. While compliance has increased as countries have experienced second waves, overall compliance has decreased somewhat since the start of the pandemic [4]. Vaccinated individuals, in particular, may feel less motivated to comply, given perceived lower health risks. Empirical evidence from the COVID-19 and previous epidemics [5–7], and predictions from influential models of health behaviour, such as the Risk Compensation, Health Belief and COM-B models [8–10], suggest that individuals who are less concerned about catching a virus have lower compliance. Further, in the UK, citizens have expressed difficultly keeping abreast of latest rules [11–14], due to variations in rules across areas and over time and (speculatively) due to “lockdown fatigue”. Vaccinated individuals may therefore not be aware of non-exemption from government rules.

Early evidence from vaccine roll-out in Israel and the UK finds some increase in infection rates following first vaccination [15,16], and infection rates have risen in Chile despite high vaccination rates [17]. Some have argued that this may reflect lower compliance with protective behaviours [18–20]. This is supported by survey evidence from early December 2020 that 40% of respondents intended to comply less or not comply with government guidelines following vaccination [21] and with recent evidence that a sizeable minority of vaccinated over 80s in the UK have subsequently broken household mixing rules [22]. Further, longitudinal evidence from influenza and Lyme’s disease vaccination programmes shows reduced compliance with some protective behaviours [23,24]. Yet, cross-sectional evidence inquiring about changes in behaviour following COVID-19 vaccination show more over-80s reporting *increased* compliance (8-15%) with hand-washing, face mask wearing, and social distancing rules than decreased compliance (1-2%) [22].

Given the risk of vaccinated individuals catching and transmitting the virus, understanding whether people comply less following vaccination is important for managing the pandemic [25]. Yet, there is a notable lack of rigorous research on the consequences of COVID-19 vaccination for personal protective behaviours [20]. Therefore, in this paper, we used monthly panel data from a large sample of UK adults to explore changes in compliance following vaccination.

## METHODS

### Sample

Data were drawn from the COVID-19 Social Study; a large ongoing panel study of the psychological and social experiences of over 70,000 adults (aged 18+) in the UK during the COVID-19 pandemic. The study commenced on 21st March 2020 and involves online weekly (from August 2020, monthly) data collection from participants for the duration of the COVID-19 pandemic in the UK. The study is not random and therefore is not representative of the UK population, but it does contain a heterogeneous sample. Participants were recruited using three primary approaches. First, convenience sampling was used, including promoting the study through existing networks and mailing lists (including large databases of adults who had previously consented to be involved in health research across the UK), print and digital media coverage, and social media. Second, more targeted recruitment was undertaken focusing on (i) individuals from a low-income background, (ii) individuals with no or few educational qualifications, and (iii) individuals who were unemployed. Third, the study was promoted via partnerships with third sector organisations to vulnerable groups, including adults with pre-existing mental health conditions, older adults, carers, and people experiencing domestic violence or abuse. The study was approved by the UCL Research Ethics Committee [12467/005] and all participants gave informed consent. The study protocol and user guide (which includes full details on recruitment, retention, data cleaning and sample demographics) are available at https://github.com/UCL-BSH/CSSUserGuide.

For these analyses, we focused on participants aged 89 or younger who completed the monthly survey in each of the six months between 23 September 2020 and 22 March 2021 (n = 23,287; 62.3% of individuals with data collection between these dates; 32.6% interviewed at any point). Ages are capped at age 90 in the data, so we excluded participants aged 90 or above from this analysis. Though there is slight overlap in calendar months, for brevity, below we refer to the survey waves as October, November, December, January, February and March waves, respectively. We used matching in this analysis and excluded participants with missing data on any variable used (n = 827; 3.6% of the eligible sample). This left a total sample size of 22,460.

The vaccine roll-out began in the UK on 8 December 2020. 768,000 individuals were vaccinated in England by 27 December 2020, 6.3 million by 28 January 2020 and 14.9 million by 25 February 2020 (1.4%, 11.4%, 27.0% of the population, respectively) [26]. The COVID-19 Social Study does not contain information on the date of vaccination, but given few individuals reported being vaccinated on, or shortly after, 23 December 2020, we assume that no participants were vaccinated before this date (1.32% of participants recorded vaccination on 23 December 2020). The vaccine was initially rolled out in age order, beginning with over 80 olds, then over 75s, and over 70s. Frontline health and social care workers, older adults in care homes, and clinically extremely vulnerable individuals were also offered the vaccine [27].

The period studied here coincides with the second wave of COVID-10 in the UK. There have been a several changes to government rules across this period. Supplementary Figure S1 displays the Oxford COVID-19 Government response tracker [28], a numeric summary of the severity of COVID-19 measures across time, as well as death rates and new case rates of COVID-19. Changes to government policy are described further in the Supplementary Information.

### Measures

Compliance was measured with two questionnaire items, which we analysed separately. *General compliance* was measured with a single-item question, “Are you following the recommendations from authorities to prevent the spread of Covid-19?”. Responses ranged from “1. Not at all” to “7. Very much so”. *Social distancing* was measured with a single question “When you go out or meet with others have you been maintaining social distancing?”. The responses categories ranged from “1. Yes, completely” to “4. Not at all” with an extra category for those who had not met with others or left their home in the last week. We reverse code this item so high scores indicate greater compliance and code those who did not leave them home or meet with others as the highest level of compliance (range 1-5).

### Statistical Analysis

Our analysis proceeded in three steps. First, we split our sample into three groups: individuals who first reported being vaccinated in the January wave; individuals who first reported being vaccinated in the February wave; and individuals who did not report being vaccinated by February. Second, given the rules used for roll-out of the vaccine, we used matching to obtain samples of similar individuals across the three groups. As our “treatment” variable (vaccination) had three levels, we carried out matching for each combination of two groups, obtaining three matched samples (January vs February vaccinators; February vs non-vaccinated; and January vs non-vaccinated). Observations were matched using Mahalanobis distance within a caliper of 0.25 SD in propensity scores. We used 1-to-1 matching without replacement and discarded observations outside the region of common support.

In the Mahalanobis distance step, given vaccine eligibility criteria, we matched upon age, date of interview in the December wave, whether the participant was a keyworker, and whether they had a flu vaccine in the past year (an indicator of existing health problems and willingness to accept vaccination). To estimate propensity scores, we used variables for age (natural splines with degrees of freedom 3), date of data collection in December (natural splines with degrees of freedom 3), keyworker status, previous flu vaccination, sex, *general compliance* and *social distancing* in the December wave (inputted as categorical variables), attitudes to vaccination (exploratory factor analysis of 12 items; September wave), intention to receive COVID-19 vaccination (September wave; categorical variable), whether the participant reported shielding for health reasons at any point, and number of chronic health conditions (0, 1, 2+) and whether the participant had a diagnosis for a psychiatric condition. More detail on these variables is given in the Supplementary Information. We assessed match quality as bias < 0.1 SD for each covariate, Rubin’s B < 0.25, Rubin’s R of 0.5-5, and visual inspection of the distributions for variables used in the Mahalanobis distance matching step.

In the third step, we estimated fixed effects regression models for each matched sample, separately, comparing within-person changes in compliance behaviour by wave of data collection across vaccination groups. Our model was of the form:

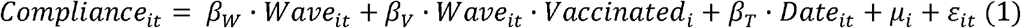

where *i* and *t* index individuals and waves respectively. *Wave*_*it*_ is a categorical variable for wave of data collection (December wave used as reference category). *Vaccinated*_*i*_ is an indicator for vaccination group; *β*_*V*_ is a vector of coefficients assessing differences in within person changes in compliance by wave of data collection; *Date*_*it*_ is a vector of date fixed effects to account for time trends in compliance behaviour; and *μ*_*i*_ and *ε*_*it*_ are person-specific and observation-specific random errors, respectively.

Our interest was in the sign and size of the coefficients *β*_*V*_. Our hypothesis was that, compared with non-vaccinated individuals, compliance would be lower among vaccinated individuals in the months that they were vaccinated, and, given that vaccination does not confer immediate immunity, progressively lower the more time had elapsed since vaccination. There should also be no differences in compliance levels in the months prior to vaccination. In our data, this hypothesis translated into no differences in compliance by vaccination status in the months October, November, and December; differences in compliance in January, February and March when comparing January vaccinators with February vaccinated or non-vaccinated individuals; and differences in compliance in February and March but not January when comparing February vaccinators with non-vaccinated individuals.

Data analysis was carried out in R v 4.0.3. [29]. Matchings was carried out using the matchit package [30], Due to stipulations set out by the ethics committee, data will be made available at the end of the pandemic. The code to replicate the analysis is available at https://osf.io/xghvb/.

## RESULTS

### Descriptive Statistics

Descriptive statistics for the full sample are displayed in Table 1. There were several differences among the vaccination groups, most notably on age, keyworker status, and date of December interview. Differences were markedly smaller following matching (Supplementary Table S1). Figures showing standardized mean differences in the study variables across matched and unmatched samples are displayed in Supplementary Figure S2-S4. Matching reduced differences in almost all cases. In the January vs February and February vs non-vaccinated comparison groups, (absolute) standardized mean differences were less than 0.1 SD in each case. The quality of the matching was lower in the January vs unvaccinated groups, though Rubin’s B and R statistics were within boundaries considered to be acceptable matching (Table 2). Supplementary Figures S5-S7 show the distributions of age, date of date collection in December, and keyworker status in the matched samples, specifically, given these are important predictors of vaccination status. Matching in the January vs February vaccination comparison group was successful, but there were notable differences in the distributions of age and survey date in the January vs non-vaccinated and February vs non-vaccinated groups, respectively.

**Table 1:**
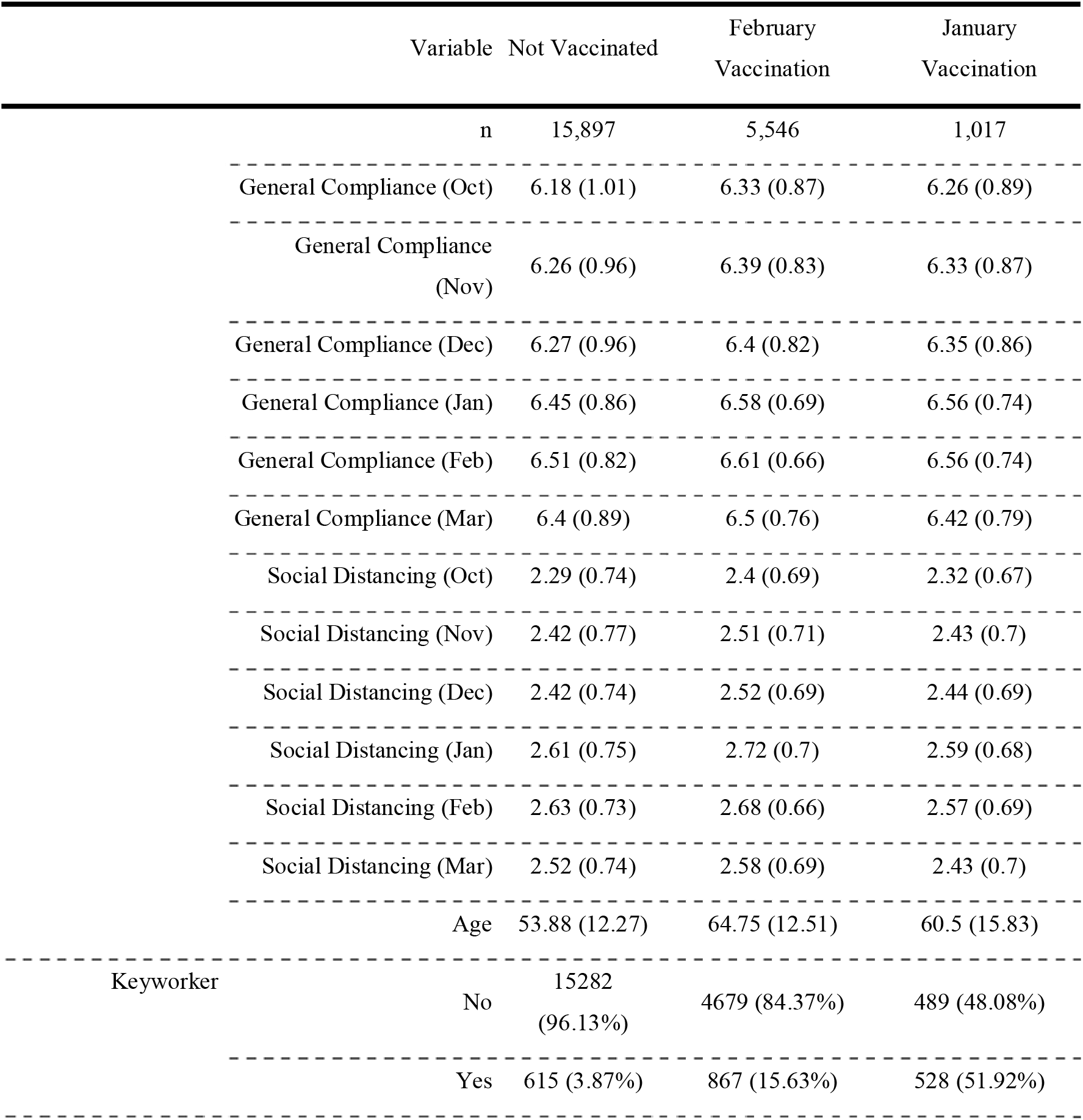
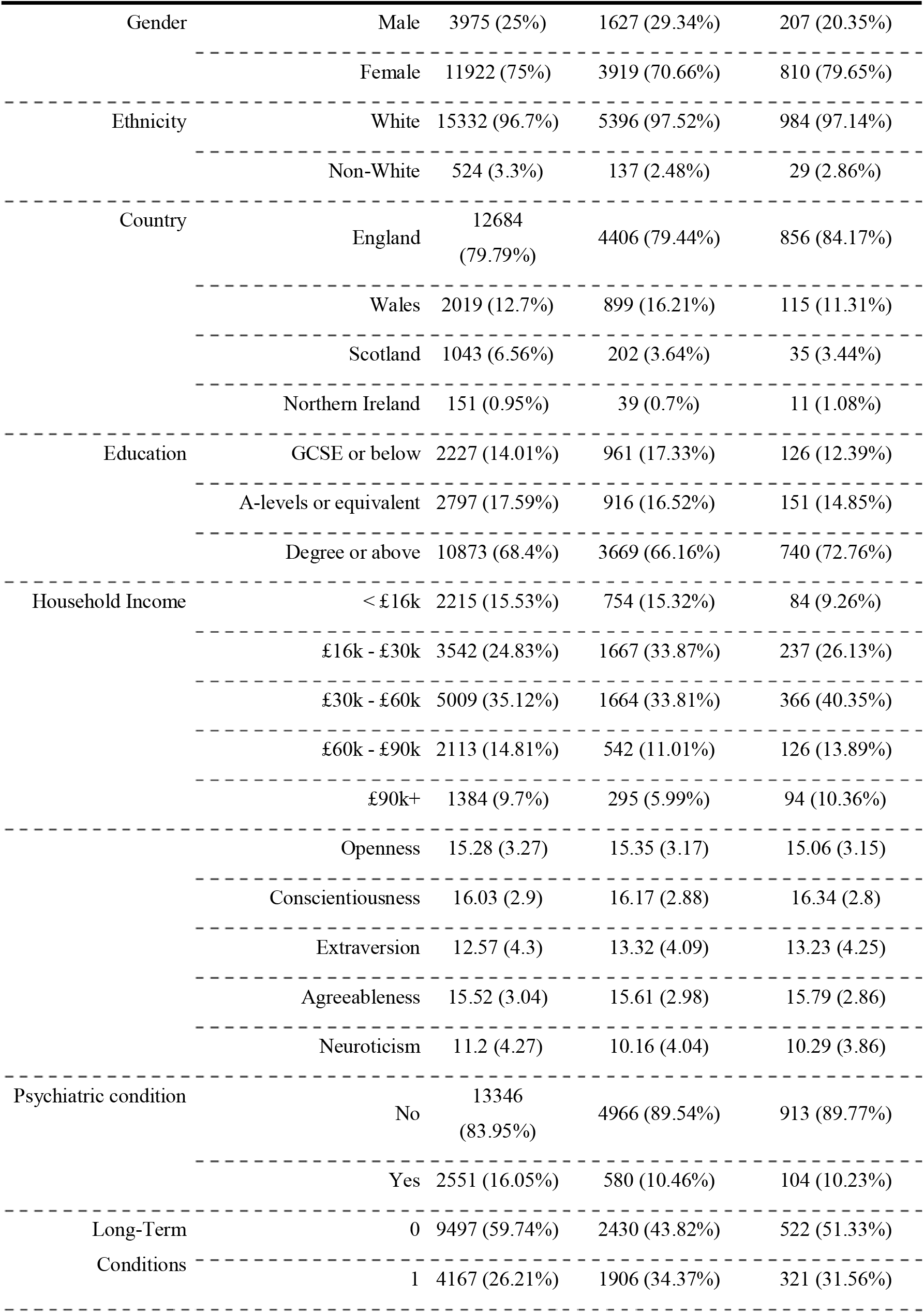
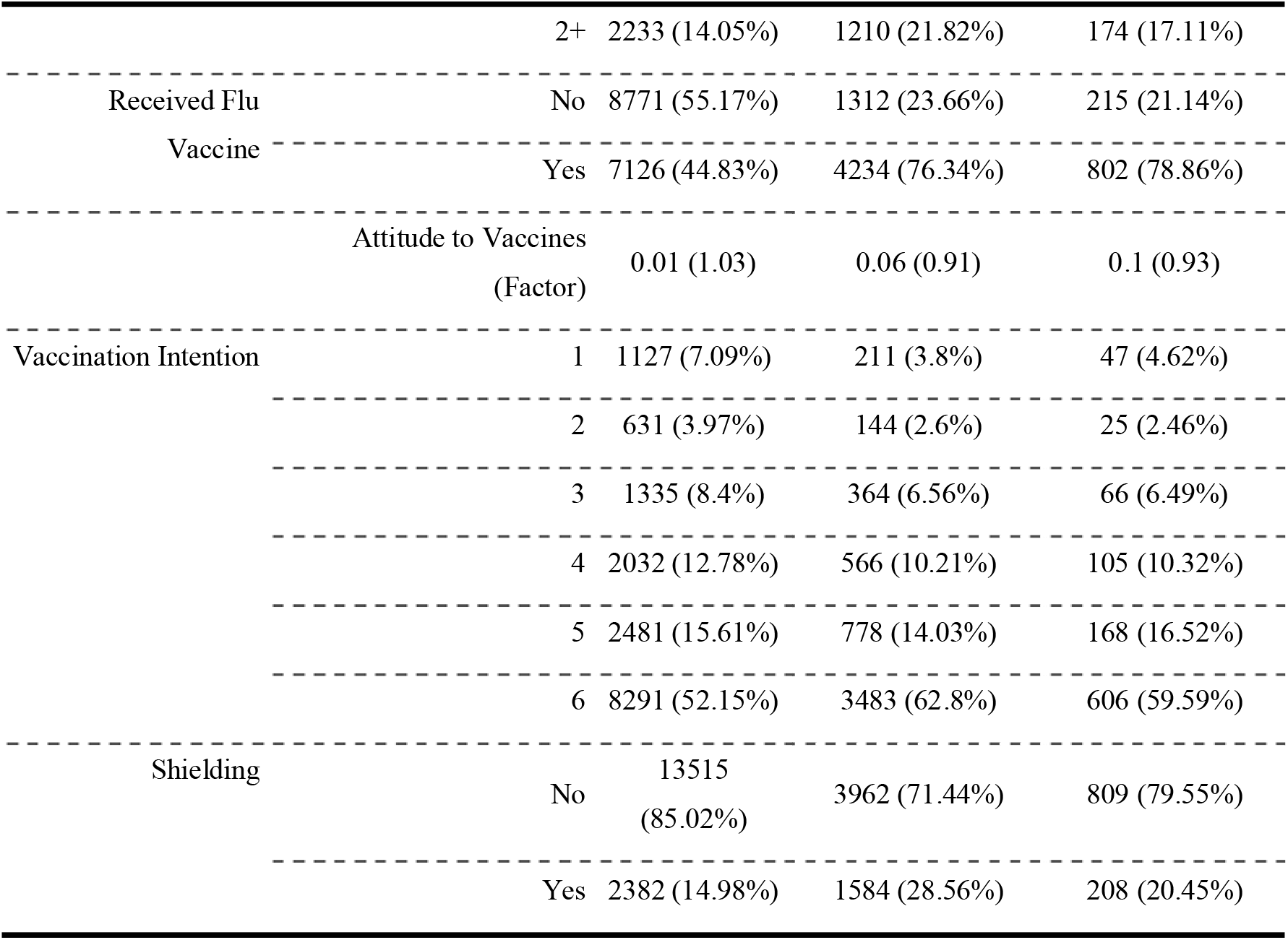
Descriptive statistics

**Table 2:** Sample sizes in matched samples

| Sample | N | Rubin's R | Rubin's B | Rubin's B (Pairs) |
| --- | --- | --- | --- | --- |
| January vs. February | 2,004 | 1.11 | 7.97 | 11.93 |
| January vs. Not Vaccinated | 1,294 | 1.11 | 6.75 | 8.23 |
| February vs. Not Vaccinated | 7,596 | 1.20 | 15.16 | 17.09 |

\* Success of the propensity score matching was assessed using Rubin's B<25%, Rubin's R of 0.5-2, and a bias of < 10% SD for each covariate.

Figure 1 shows the trends in each compliance measure over the study period. As the UK entered a second wave, there were increases in both compliance measures, though with some decrease in social distancing over December [3,see also 31].

**Figure 1:**
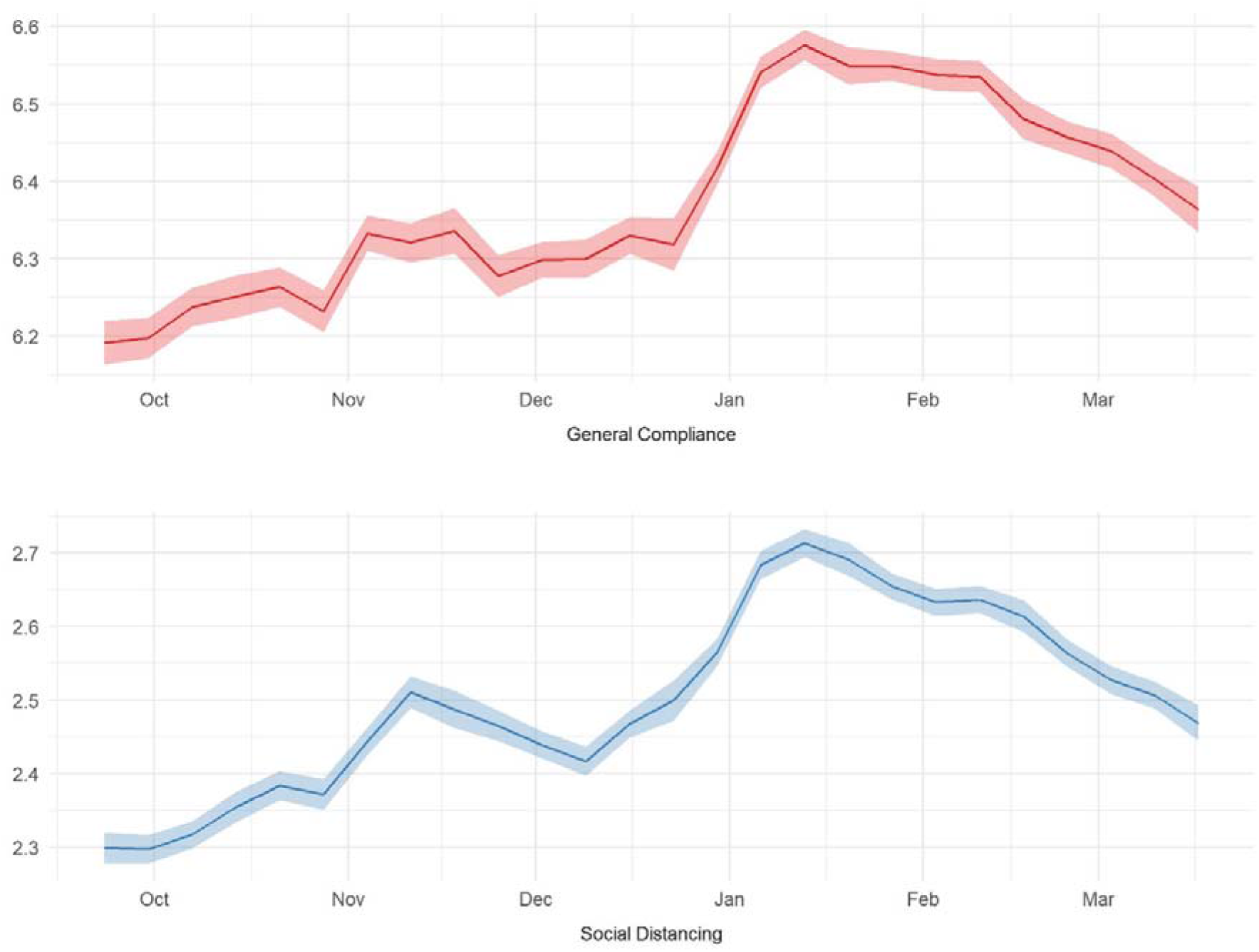
Trends in compliance behaviours

### Vaccinations and Compliance Behaviour

The results of the fixed effects regressions are displayed in Figure 2. There were no statistically significant differences in either compliance measure following vaccination in any matched sample group. There were also no statistical significant differences prior to vaccination, suggesting this in no biased by unobserved confounding in the matched samples.

**Figure 2:**
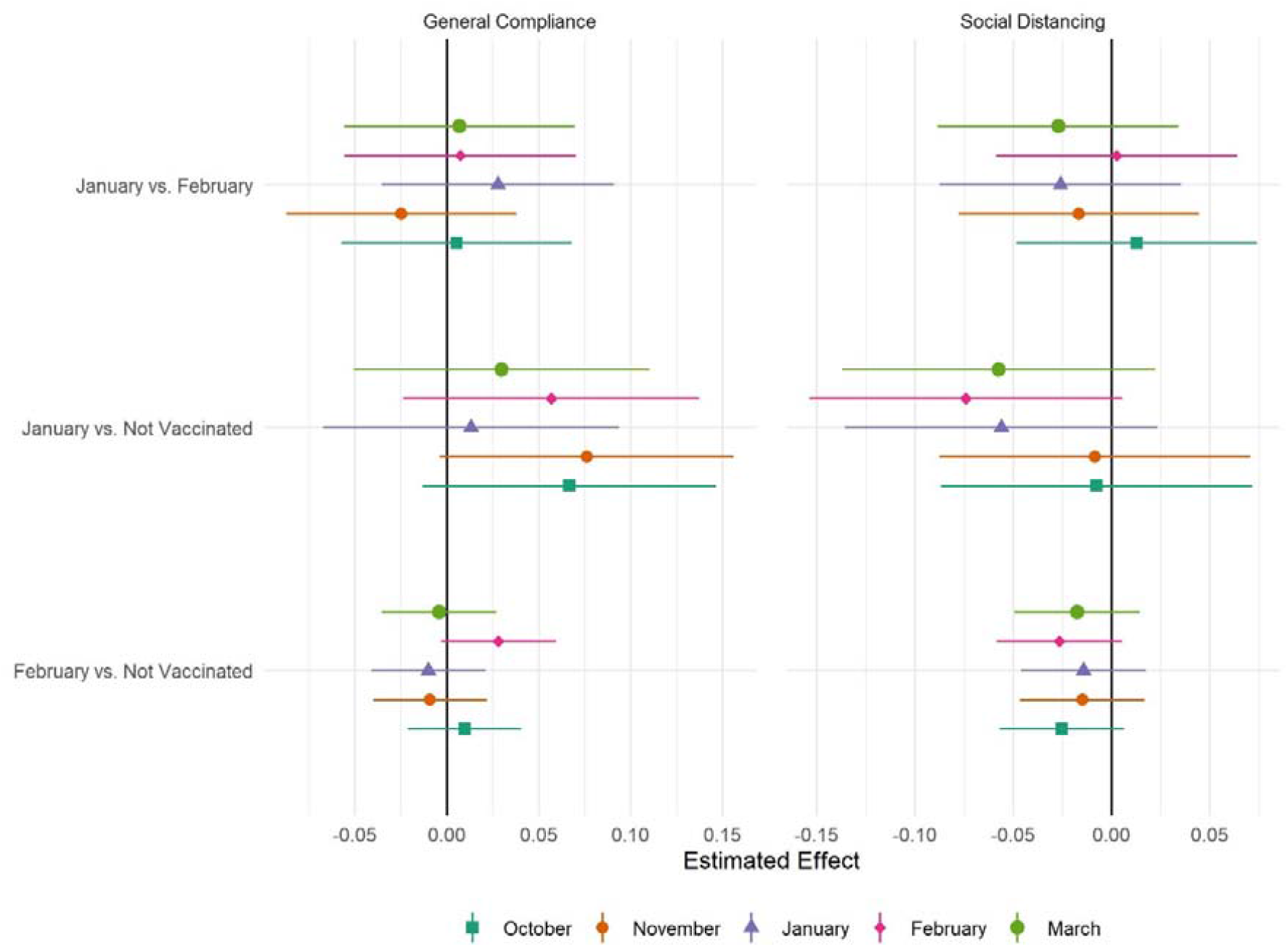
Results of fixed effects regression by matched sample and measure of compliance

It is possible that small average differences may mask heterogeneous effects – a small number of vaccinated individuals could stop complying altogether. To explore this, Figure 3 displays bar plots for compliance levels at each interview in the January vs February vaccination matched sample. There was no clear evidence of extremely low levels of compliance in the vaccinated group. The same is true when comparing February vaccinators or January vaccinators with non-vaccinated individuals (Supplementary Figures S8 and S9). In fact, as shown in Supplementary Table S1, average compliance levels increased among all groups between October and February in line with the increase in compliance seen in the wider population.

**Figure 3:**
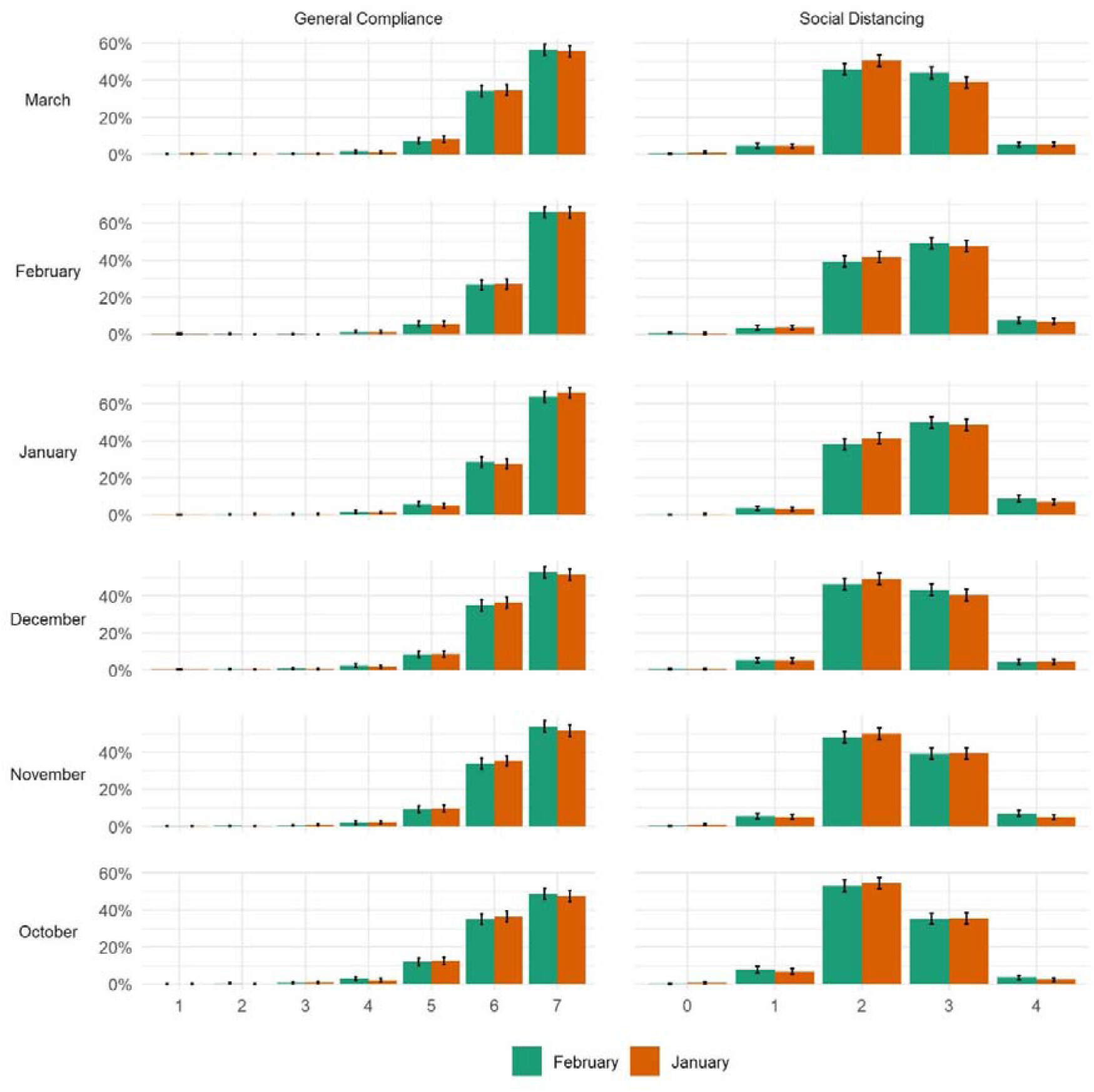
Distribution of compliance behaviours by vaccination status and wave, January vs February vaccinated matched sample.

### Sensitivity Analyses

Given that fixed effects regressions compare within-person changes in compliance levels across vaccination groups, we also repeated the model in (1) using mixed effects modelling, interpreting the term *u*_*i*_ as a normally-distributed random intercept. These regressions tested differences in compliance *levels* by vaccination status and wave. The results are shown in Supplementary Figure S10 and are qualitatively similar to those shows in Figure 2.

## DISCUSSION

Using panel data from five months of the pandemic in the UK, we found no clear evidence that receiving a COVID-19 vaccine reduced compliance behaviour. Descriptively, there was little evidence of vaccinated individuals reducing compliance altogether. In fact, vaccinated individuals – like non-vaccinated individuals – increased compliance from the beginning of the period as the UK experienced its second wave of COVID-19.

The results are striking given existing evidence that compliance levels are higher among those with greater health risks from – or greater fears of – catching COVID-19 [5,7], and evidence of widespread intentions to reduce compliance following vaccination [21]. An explanation for the discrepancy may be the almost exclusive use of cross-sectional data in the literature – a recent study shows that marked differences in between-person and within-person associations between compliance and several factors [32]. The results suggest that vaccinations do not crowd-out other preventive behaviours. However, it should be noted that we used a relatively short follow-up period – differences in compliance may take time to arise, especially as individuals are warned that vaccines do not take effect immediately and second vaccinations are required for full effectiveness. Vaccinated individuals in our sample were also relatively old. The results may have been different were vaccinations rolled out more widely. For instance, intentions to reduce compliance or not comply following vaccination are higher among younger age groups [21]. Further, compliance was measured during a period of strict lockdown where the opportunities for non-compliance were limited. This study should be repeated as lockdowns are eased. We also only focused on two measures of compliance. Differences could potentially be observed for other behaviours, such as indoor or outdoor household mixing.

This study had a number of other limitations. First, we used two self-report measures of compliance which may be subject to biases such as recall bias or social desirability bias. Being vaccinated could be considered a form of compliance so our general compliance measure may not have been specific enough to pick up on differences in specific compliance behaviour. Second, our sample was not representative and, moreover, comprised of individuals who comply more than on average [33]. This may have biased associations toward the null. Third, the existence of the vaccine program may have induced behaviour changes in the non-vaccinated group, if these individuals were less concerned about transmitting the virus [34]. Fourth, compliance was changing over time, even in the absence of vaccination. Previous research has shown that the strength of several factors in predicting compliance differs over pandemics [33,35]. Our matched samples may therefore not provide an appropriate counterfactual and results may be biased by unobserved confounding. Nevertheless, by exploiting the longitudinal nature of our sample, we were able to use compliance in months prior to vaccination as a placebo test. No statistically significant differences were found in these months, which may add confidence to our results.

Our results suggest that there is no immediate cause for concern of widespread non-compliance among vaccinated individuals. However, it is important to continue monitoring the situation as the vaccine is roll-out more widely, restrictions are lifted, and people receive second doses. Analyses using data from other populations and that examine the potential impact of widespread vaccination on the behaviour of those not yet vaccinated are also required in order to ensure that the gains of the vaccination program are not lost through increases in risky behaviour.

## Data Availability

Data used in this study will be made publicly available once the pandemic is over. The code used to run the analysis is available at https://osf.io/xghvb/.

## Acknowledgements

The researchers are grateful for the support of a number of organisations with their recruitment efforts including: the UKRI Mental Health Networks, Find Out Now, UCL BioResource, HealthWise Wales, SEO Works, FieldworkHub, and Optimal Workshop.

## Author contributions

LW, AS, HWM, and DF conceived and designed the study. LW analysed the data and wrote the first draft. All authors provided critical revisions.

## Patient and Public Involvement

The research questions in the UCL COVID-19 Social Study built on patient and public involvement as part of the UKRI MARCH Mental Health Research Network, which focuses on social, cultural and community engagement and mental health. This highlighted priority research questions and measures for this study. Patients and the public were additionally involved in the recruitment of participants to the study and are actively involved in plans for the dissemination of findings from the study.

