## Supplemental Information for "Do people reduce compliance with COVID-19 guidelines following vaccination? A longitudinal analysis of matched UK adults"

SUPPLEMENTARY INFORMATION

### POLICY CONTEXT

The period studied here coincides with the (ongoing) second wave of COVID-10 in the UK. There have been several changes to government rules across this period. Figure S1 displays the Oxford COVID-19 Government response tracker ^1^, a numeric summary of the severity of COVID-19 measures across time, as well as death rates and new case rates of COVID-19. Management of the pandemic has been devolved at the nation level of England, Scotland, Northern Ireland and Wales. On 23 September, pubs and restaurants were open and households were allowed to mix indoors, though the UK’s COVID-19 alert level was raised to “high or increasing exponentially”. On 1 October, household mixing indoors was banned in North-East. On 12 October, a three-tier framework was introduced in England with Liverpool city region plan in the highest tier, Tier 3, leading to the closure of pubs. Several regions were upgraded to higher tiers in the following weeks. On 21 October, Wales entered a three-week lockdown, and on 5 November, a four-week national lockdown in England came into force. A new, stricter three-tier system was introduced in England on 2 December, with a fourth tier introduced from 19 December introduced in parts of South East England due to the spread of a new variant of the COVID-19 virus, in effect a new lockdown. Restrictions were also tightened in Wales and Scotland entered into the highest tier of its own tier system, similar to the Tier 4 in England. A relaxation in restrictions was allowed in most parts of the UK on Christmas day. On 30 December, the whole of England was placed into Tier 4. On 5 January 2021, a third national lockdown began.

### MEASURES

Details on the individual measures used for matching are below.

#### Demographics and socio-economic position

We included demographic variables for sex (male or female), age, and whether the participant was employed in a keyworker role. Keyworking was defined as working in health, social care or support sectors, or work involving in medicines or PPE production or distribution. Each was measured at baseline data collection. We also included date of data collection in December wave as a matching variable. This variable is associated with compliance behaviour (see Figure 1) and is highly correlated with dates of data collection in later months as individuals were emailed one month after their prior data collection.

#### Compliance and Confidence in Government

*General compliance* and *Social distancing* in the December wave were included as categorical variables. *General compliance* was measured with a single-item question, “Are you following the recommendations from authorities to prevent the spread of Covid-19?”. Responses ranged from “1. Not at all” to “7. Very much so”. *Social distancing* was measured with a single question “When you go out or meet with others have you been maintaining social distancing?”. The responses categories ranged from “1. Yes, completely” to “4. Not at all” with an extra category for those who had not met with others or left their home in the last week. We reverse coded this item so high scores indicated greater compliance and code those who did not leave them home or meet with others as the highest level of compliance (range 1-5).

#### Health

Long-term conditions was measured using a categorical variables (0, 1, 2+). We derived the variables by from a multiple choice question on diagnosed health conditions. The listed categories were: high blood pressure, diabetes, heart disease, lung disease, cancer, other clinically diagnosed chronic health condition, disability that affects ability to leave house, and any other disability. This question was asked at baseline interview.

Diagnosed mental health condition was measured using a categorical variable (yes, no) derived from the same multiple choice question. The relevant categories were: clinically diagnosed depression, clinically-diagnosed anxiety, and any other clinically diagnosed mental health problem.

We measured shielding as reporting not leaving home at all due to an existing medical condition or being categorised as high risk.  Survey items on shielding were included in all data collections from 12 April – 04 July 2020. We defined a person as shielding if they stated shielding at any data collection.

#### Vaccination Attitudes and Experiences

Questions of vaccination attitudes, plans, and experiences were collected between 07 September and 15 October. Attitude to vaccines was measured using the 12-item Vaccination Attitudes Examination scale (VAX) ^2^. Participants were asked to focus on vaccines in general rather than specifically on vaccines for COVID-19. Response options ranged from 1 “strongly agree” to 6 “strongly disagree.” Items are displayed in table below. We use exploratory factor analysis with polychoric correlations to extract a single factor from this measure (eigenvalue = 6.65).

Item

1. I feel safe after being vaccinated
2. I can rely on vaccines to stop serious infectious diseases
3. I feel protected after getting vaccinated
4. Although most vaccines appear to be safe, there may be problems that we have not yet discovered
5. Vaccines can cause unforeseen problems in children
6. I worry about the unknown effects of vaccines in the future
7. Vaccines make a lot of money for pharmaceutical companies, but do not do much for regular people
8. Authorities promote vaccination for financial gain, not for people's health
9. Vaccination programs are a big con
10. Natural immunity lasts longer than a vaccination
11. Natural exposure to viruses and germs gives the safest protection
12. Being exposed to diseases naturally is safer for the immune system than being exposed through vaccination

COVID-19 vaccination plans were collected using a single item measure: “How likely to do you think you are to get a COVID-19 vaccine when one is approved?”. Items were score on a 1 (“Very unlikely” to 7 (“Very likely”) measure. This item was measured between 07 September – 15 October 2020. Whether the participant had had a flu vaccination in the last year collected using a single-item question asked between the same dates.

#### Additional Variables

We tried including several other measures to improve our matching procedure, including personality traits (such as Big-5 personality traits, locus of control, and risk-taking behaviour), ethnicity (White, Non-White), and having previously refused a vaccine. The quality of the matching was worse when these variables were included and were not considered in the analysis.

### FIGURES

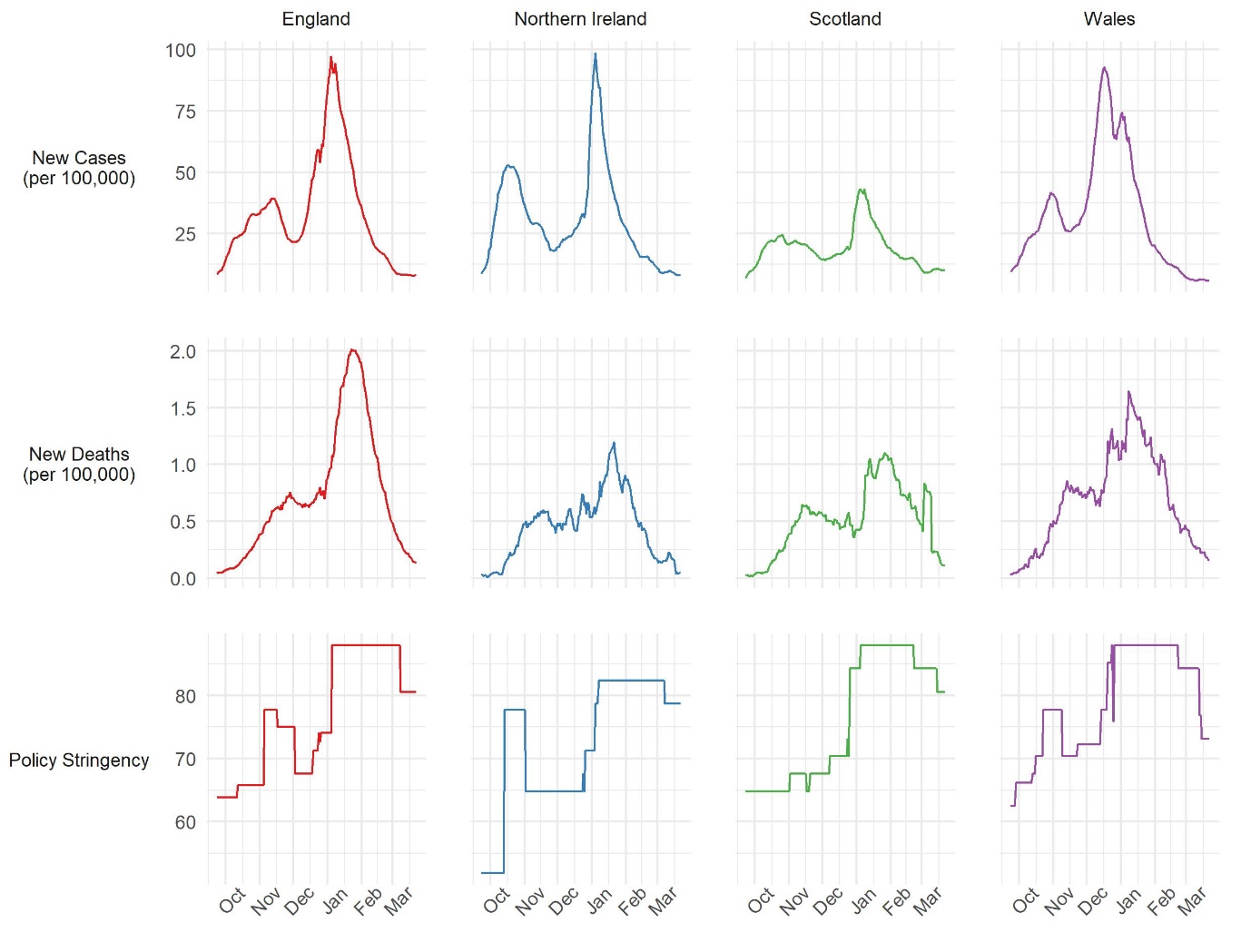

Figure S1: Oxford COVID-19 Government Response Tracker, (rolling weekly average) daily COVID-19 caseloads and deaths from COVID-19. Source: Hale et al. ^1^

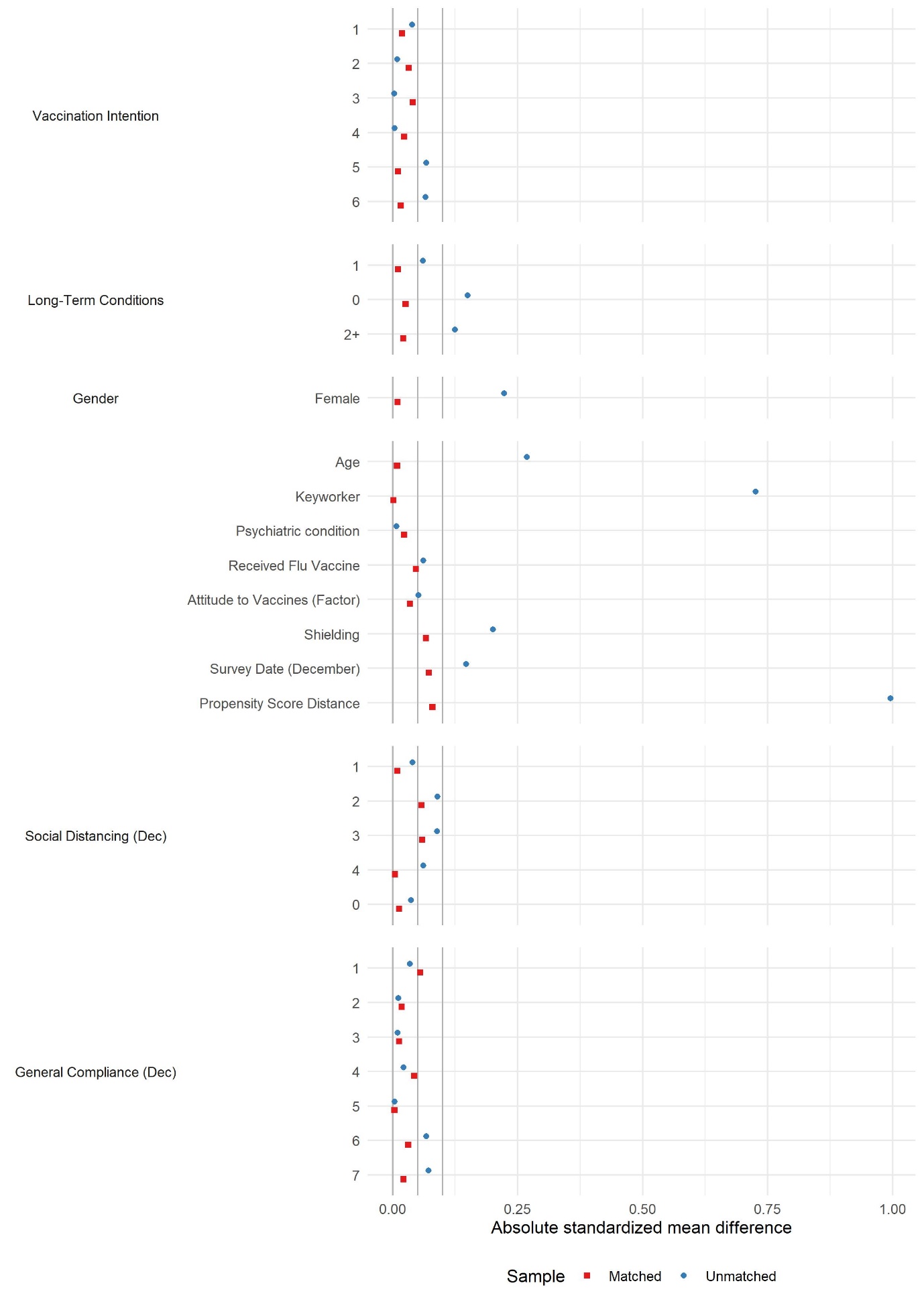

Figure S2: Absolute standardised mean differences in unmatched and matched samples, January vs February vaccinators. Vertical lines at 0, 0.05 and 0.10.

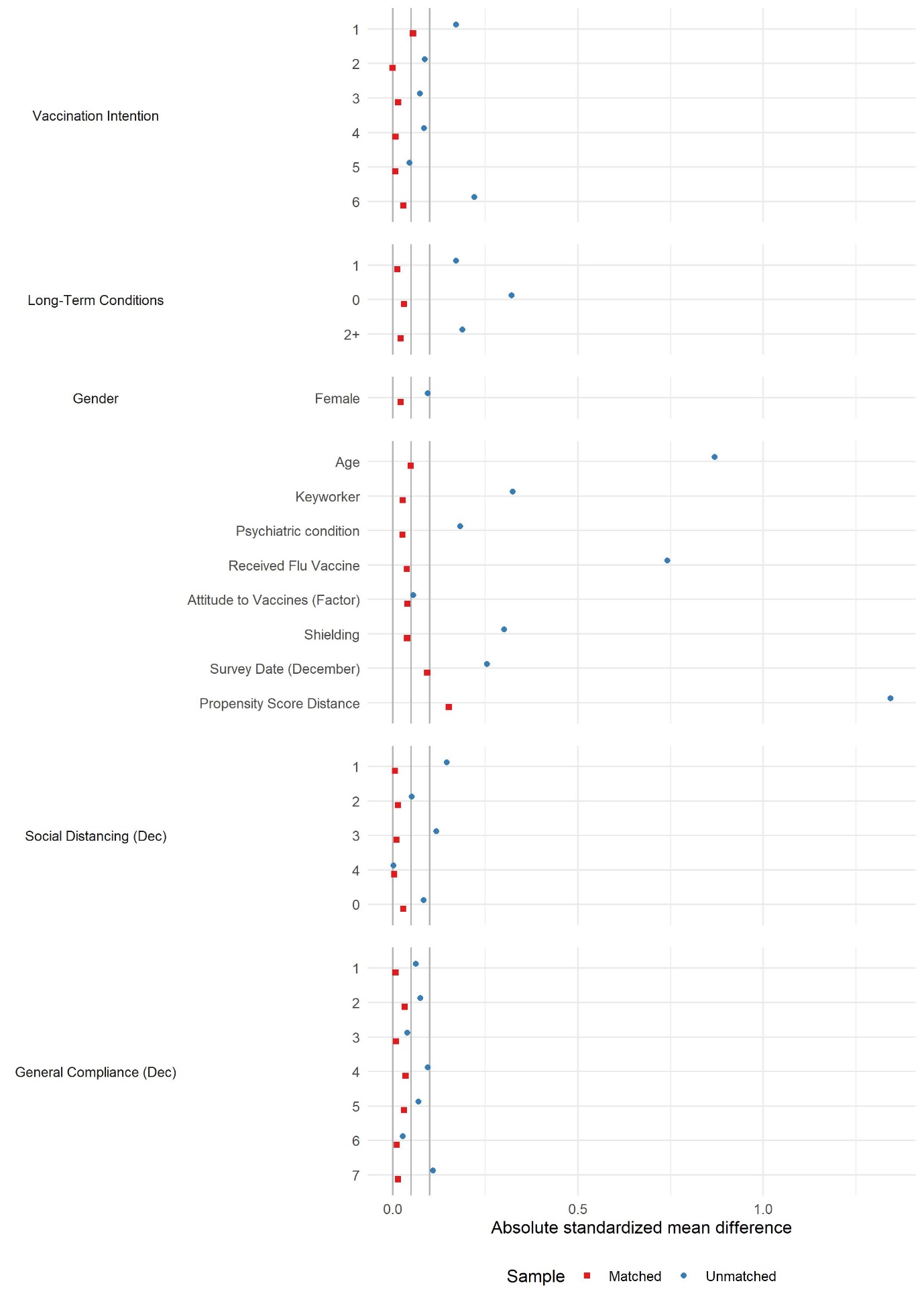

Figure S3: Absolute standardised mean differences in unmatched and matched samples, February vaccinators vs unvaccinated. Vertical lines at 0, 0.05 and 0.10.

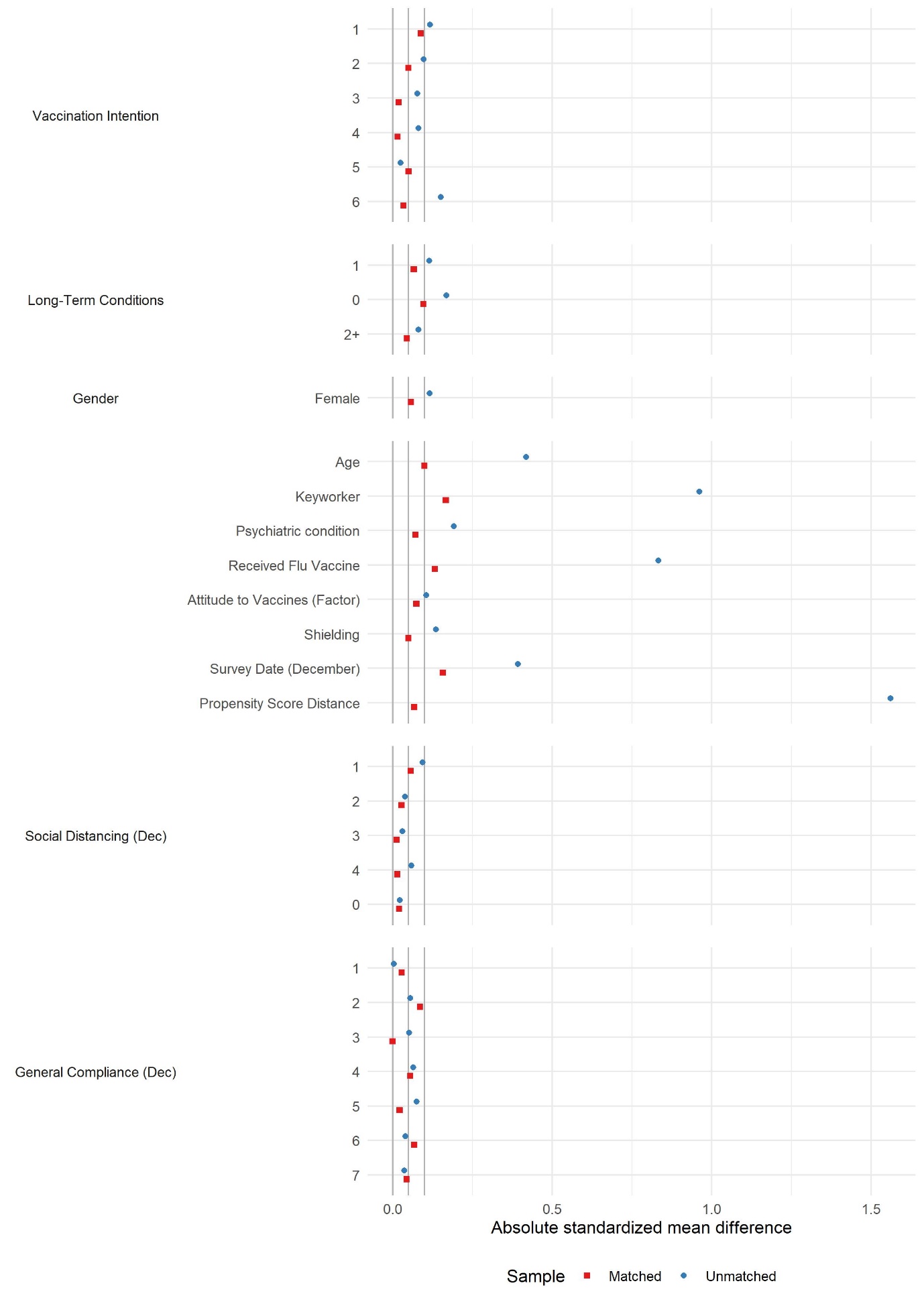

Figure S4: Absolute standardised mean differences in unmatched and matched samples, January vaccinators vs unvaccinated. Vertical lines at 0, 0.05 and 0.10.

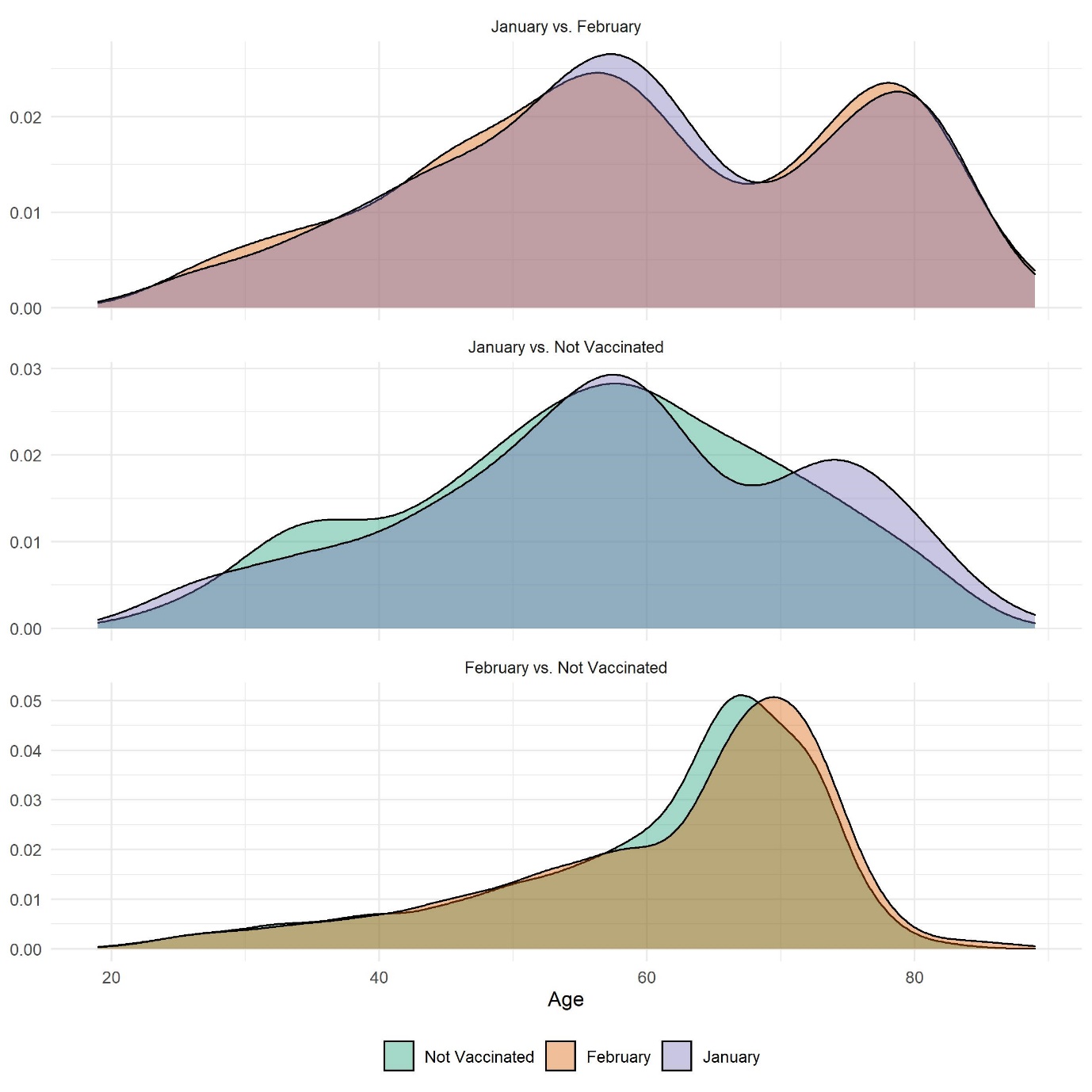

Figure S5: Distribution of age in matched sample by vaccination group and matched sample.

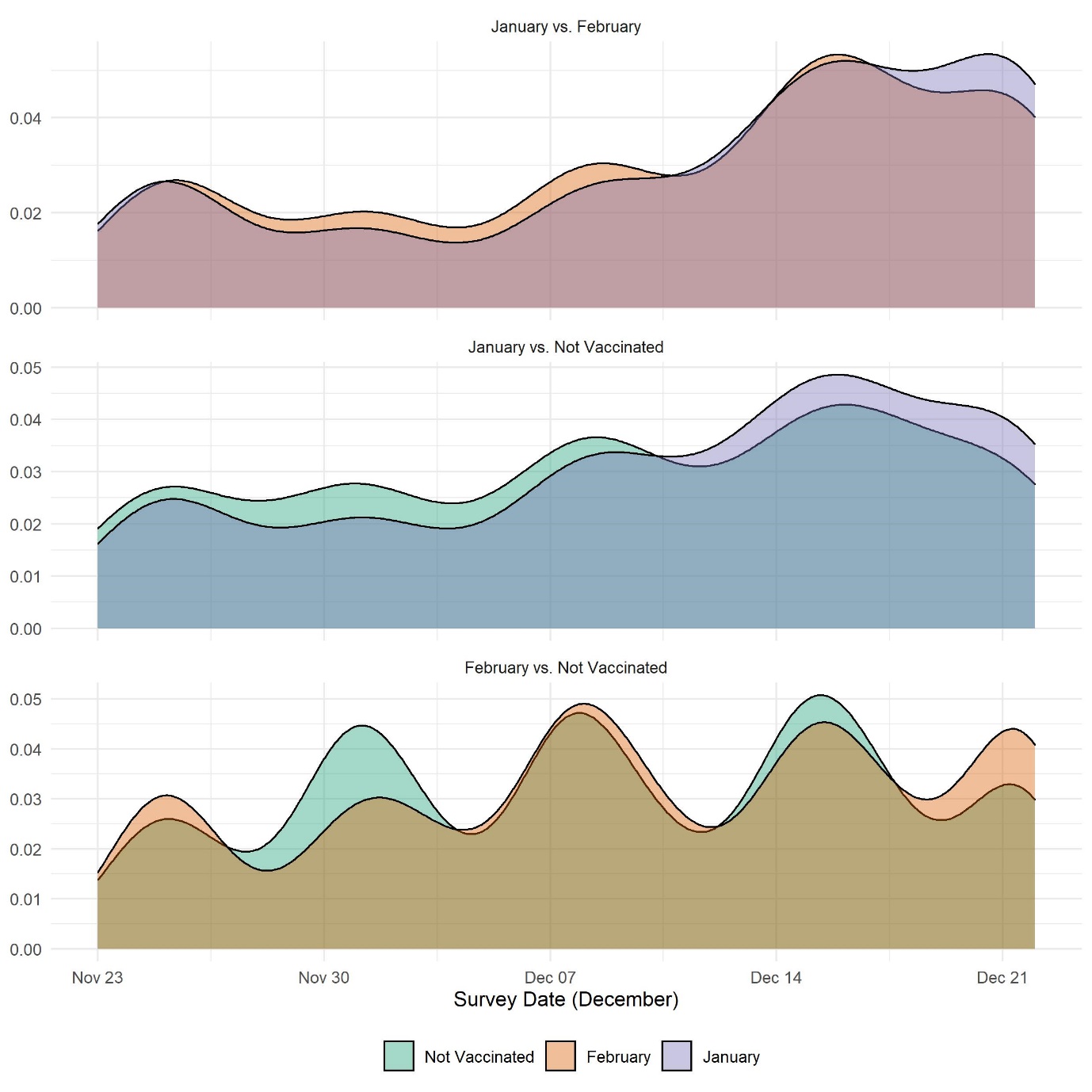

Figure S6: Distribution of date of data collection in December by vaccination group and matched sample

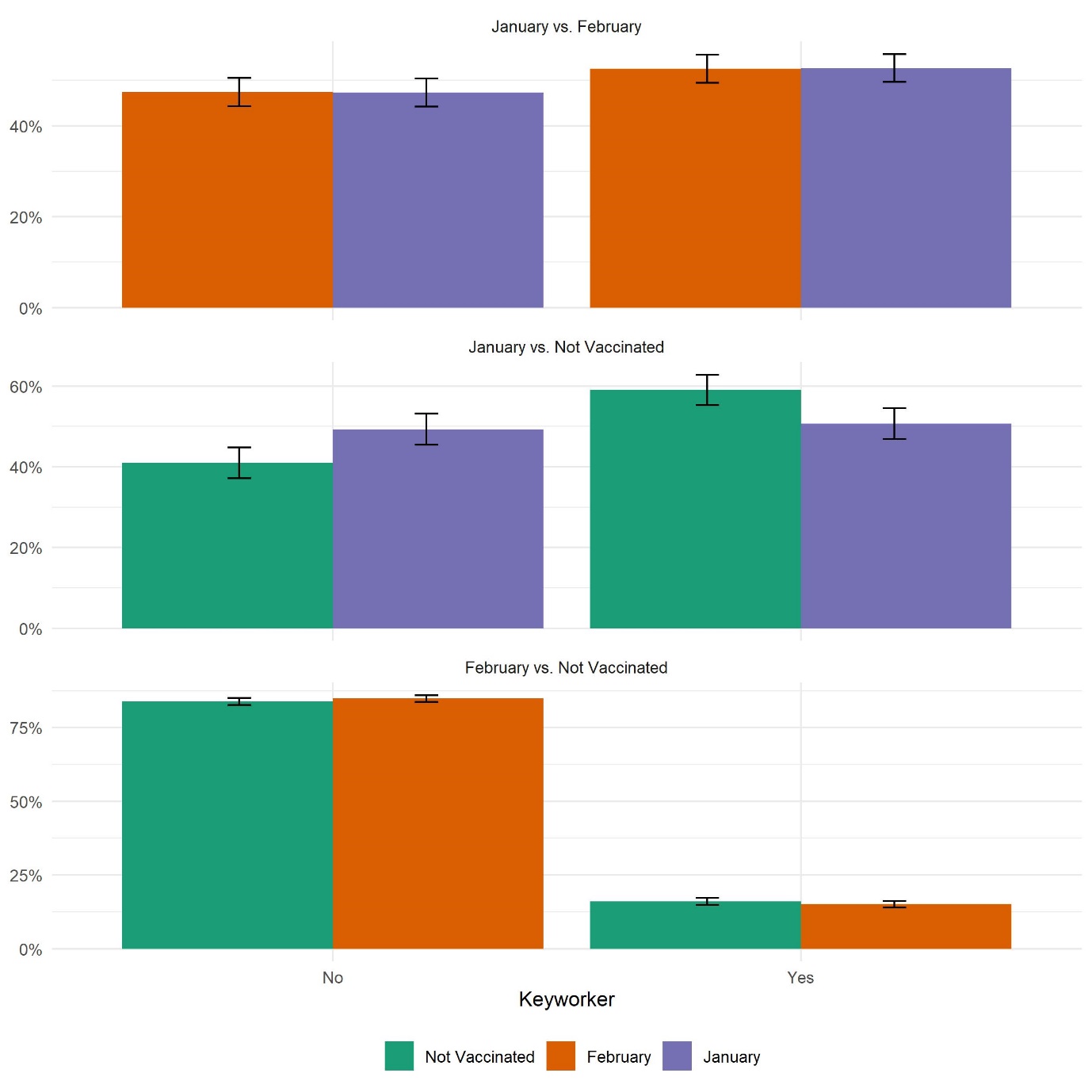

Figure S7: Distribution of keyworker status in December by vaccination group and matched sample

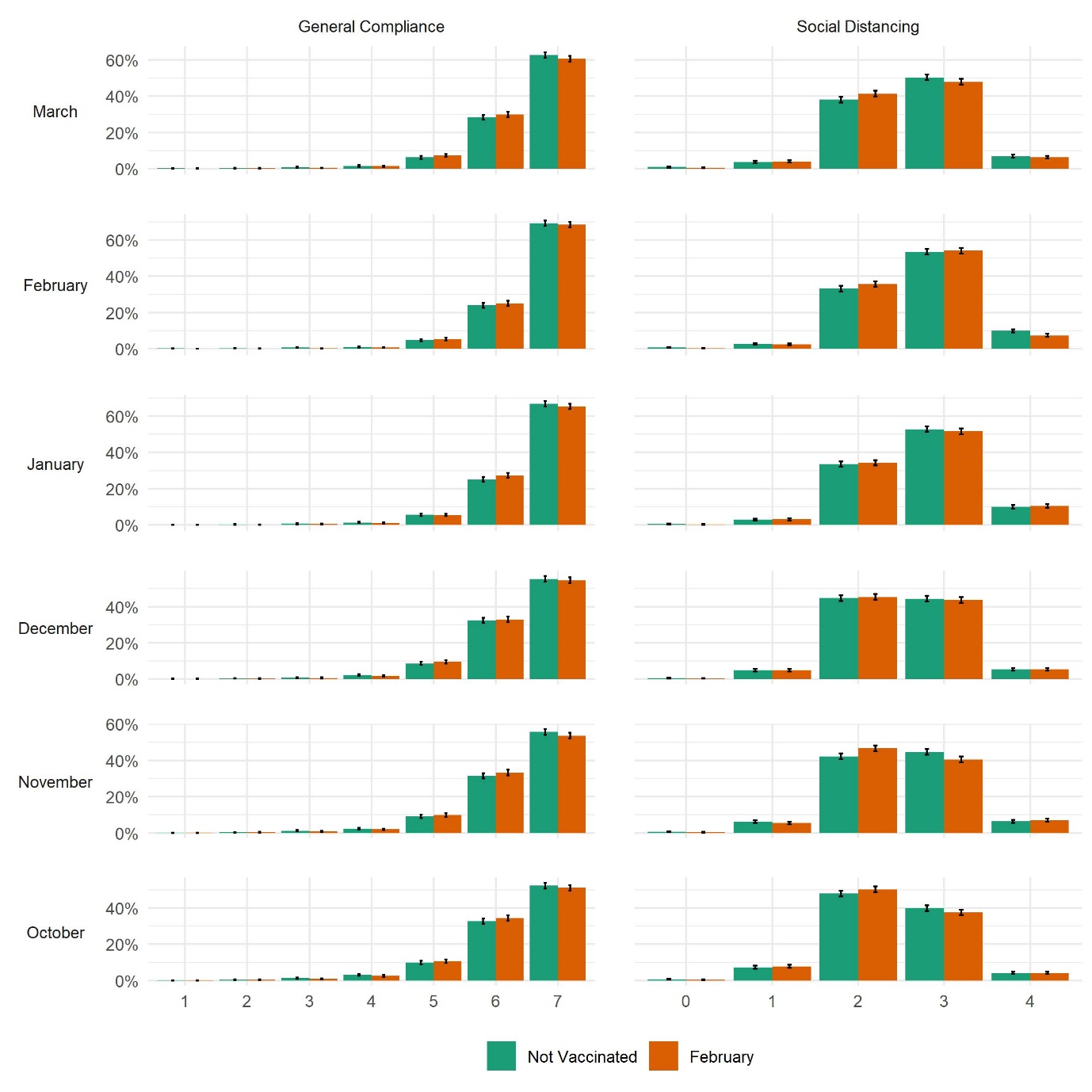

Figure S8: Distribution of compliance behaviours by vaccination status and wave, February vs non-vaccinated matched sample

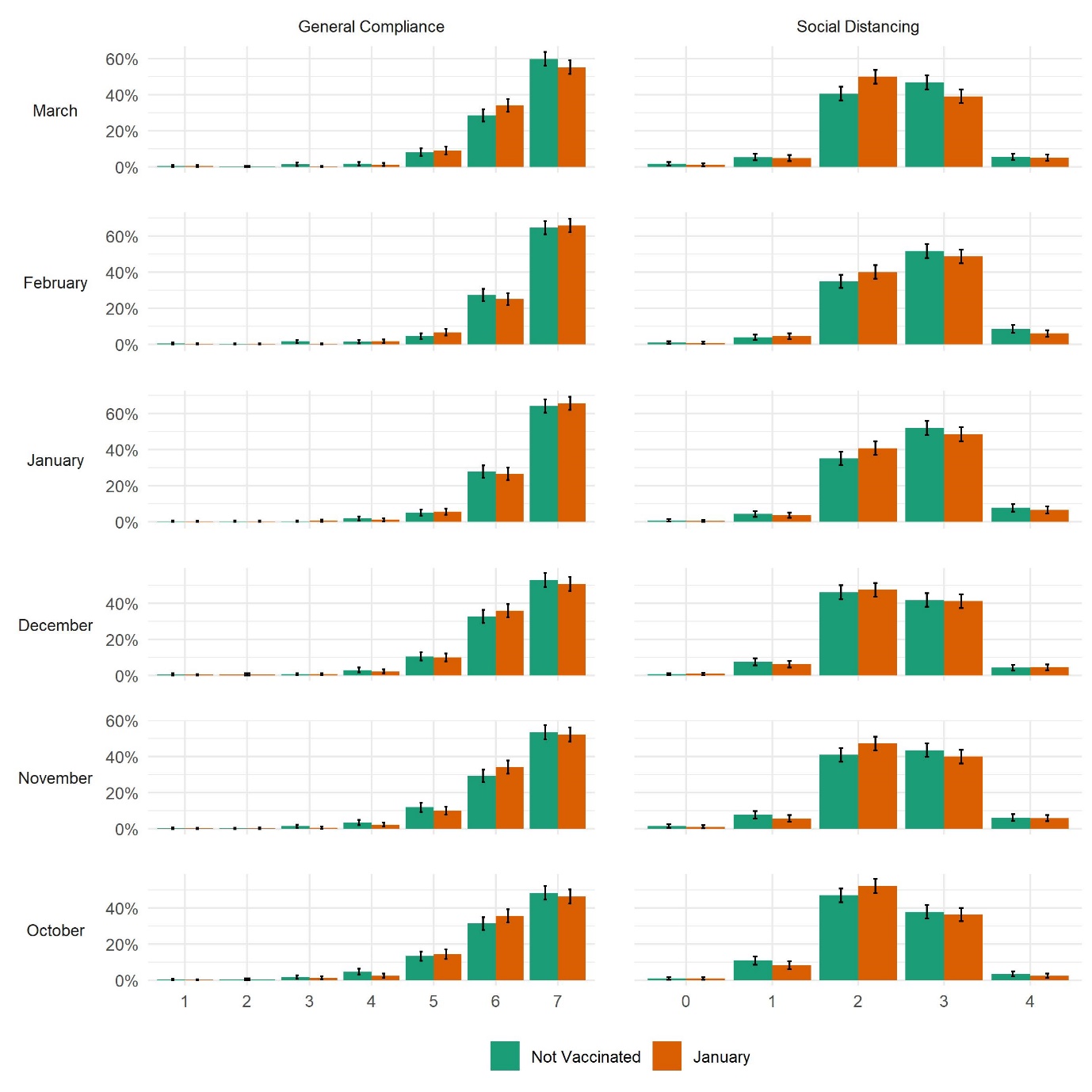

Figure S9: Distribution of compliance behaviours by vaccination status and wave, January vs non-vaccinated matched sample

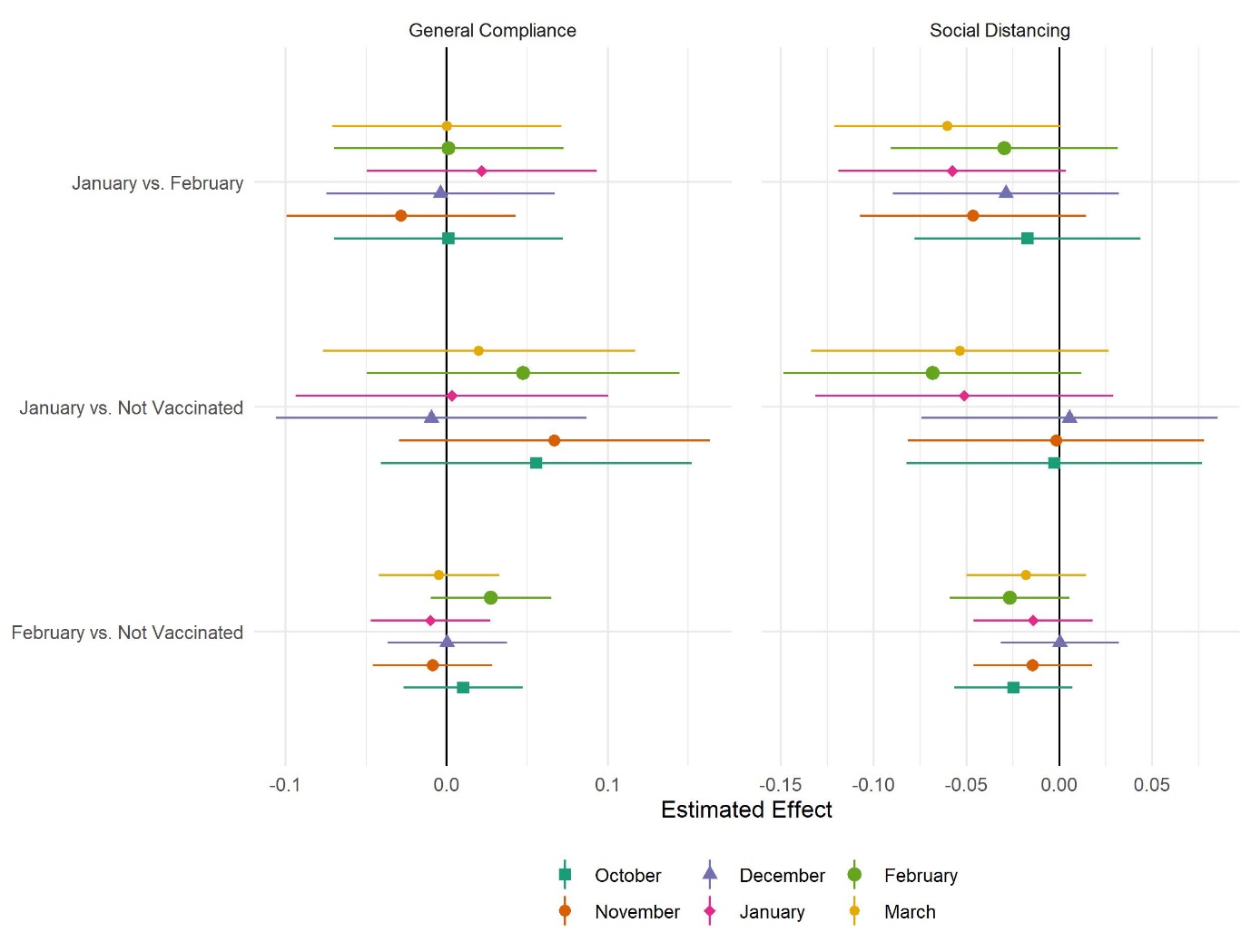

Figure S10: Results of mixed effect regressions by compliance measure and matched sample.

### TABLES

Table S1: Descriptive statistics in the matched samples

|  | | January vs. February | | January vs. Not Vaccinated | | February vs. Not Vaccinated | |
| --- | --- | --- | --- | --- | --- | --- | --- |
|  | Variable | February | January | Not Vaccinated | January | Not Vaccinated | February |
|  | n | 1,002 | 1,002 | 647 | 647 | 3,798 | 3,798 |
|  | General Compliance (Oct) | 6.26 (0.9) | 6.26 (0.89) | 6.18 (1.03) | 6.23 (0.89) | 6.3 (0.95) | 6.3 (0.9) |
|  | General Compliance (Nov) | 6.37 (0.83) | 6.34 (0.86) | 6.28 (0.97) | 6.34 (0.86) | 6.37 (0.88) | 6.36 (0.85) |
|  | General Compliance (Dec) | 6.36 (0.85) | 6.35 (0.86) | 6.32 (0.91) | 6.31 (0.9) | 6.37 (0.87) | 6.38 (0.84) |
|  | General Compliance (Jan) | 6.53 (0.73) | 6.56 (0.73) | 6.52 (0.81) | 6.54 (0.78) | 6.55 (0.77) | 6.56 (0.72) |
|  | General Compliance (Feb) | 6.56 (0.72) | 6.56 (0.74) | 6.5 (0.88) | 6.53 (0.79) | 6.58 (0.77) | 6.6 (0.67) |
|  | General Compliance (Mar) | 6.44 (0.78) | 6.43 (0.79) | 6.41 (0.92) | 6.41 (0.81) | 6.49 (0.81) | 6.48 (0.78) |
|  | Social Distancing (Oct) | 2.34 (0.69) | 2.32 (0.67) | 2.32 (0.75) | 2.31 (0.69) | 2.4 (0.71) | 2.37 (0.7) |
|  | Social Distancing (Nov) | 2.47 (0.72) | 2.42 (0.7) | 2.45 (0.79) | 2.44 (0.74) | 2.5 (0.73) | 2.48 (0.72) |
|  | Social Distancing (Dec) | 2.46 (0.69) | 2.43 (0.69) | 2.41 (0.72) | 2.42 (0.71) | 2.49 (0.7) | 2.49 (0.7) |
|  | Social Distancing (Jan) | 2.63 (0.69) | 2.59 (0.68) | 2.62 (0.72) | 2.57 (0.69) | 2.69 (0.71) | 2.69 (0.71) |
|  | Social Distancing (Feb) | 2.6 (0.7) | 2.57 (0.69) | 2.63 (0.73) | 2.55 (0.71) | 2.69 (0.71) | 2.66 (0.66) |
|  | Social Distancing (Mar) | 2.49 (0.68) | 2.43 (0.7) | 2.49 (0.76) | 2.42 (0.71) | 2.59 (0.71) | 2.56 (0.69) |
|  | Age | 59.96 (15.93) | 60.1 (15.6) | 56.06 (13.93) | 57.64 (14.75) | 61.15 (12.01) | 61.75 (12.39) |
| Keyworker | No | 475 (47.41%) | 474 (47.31%) | 265 (40.96%) | 319 (49.3%) | 3186 (83.89%) | 3224 (84.89%) |
|  | Yes | 527 (52.59%) | 528 (52.69%) | 382 (59.04%) | 328 (50.7%) | 612 (16.11%) | 574 (15.11%) |
| Gender | Male | 195 (19.46%) | 199 (19.86%) | 109 (16.85%) | 124 (19.17%) | 1017 (26.78%) | 1054 (27.75%) |
|  | Female | 807 (80.54%) | 803 (80.14%) | 538 (83.15%) | 523 (80.83%) | 2781 (73.22%) | 2744 (72.25%) |
| Ethnicity | White | 965 (96.6%) | 970 (97.19%) | 623 (96.44%) | 624 (96.74%) | 3697 (97.65%) | 3688 (97.18%) |
|  | Non-White | 34 (3.4%) | 28 (2.81%) | 23 (3.56%) | 21 (3.26%) | 89 (2.35%) | 107 (2.82%) |
| Country | England | 759 (75.75%) | 842 (84.03%) | 486 (75.12%) | 535 (82.69%) | 2820 (74.25%) | 3117 (82.07%) |
|  | Wales | 186 (18.56%) | 115 (11.48%) | 102 (15.77%) | 83 (12.83%) | 701 (18.46%) | 517 (13.61%) |
|  | Scotland | 49 (4.89%) | 34 (3.39%) | 54 (8.35%) | 20 (3.09%) | 232 (6.11%) | 134 (3.53%) |
|  | Northern Ireland | 8 (0.8%) | 11 (1.1%) | 5 (0.77%) | 9 (1.39%) | 45 (1.18%) | 30 (0.79%) |
| Education | GCSE or below | 133 (13.27%) | 124 (12.38%) | 87 (13.45%) | 70 (10.82%) | 643 (16.93%) | 637 (16.77%) |
|  | A-levels or equivalent | 127 (12.67%) | 149 (14.87%) | 104 (16.07%) | 100 (15.46%) | 663 (17.46%) | 647 (17.04%) |
|  | Degree or above | 742 (74.05%) | 729 (72.75%) | 456 (70.48%) | 477 (73.72%) | 2492 (65.61%) | 2514 (66.19%) |
| Household Income | < £16k | 111 (12.27%) | 82 (9.16%) | 87 (14.8%) | 46 (7.92%) | 615 (18.29%) | 497 (14.63%) |
|  | £16k - £30k | 264 (29.17%) | 234 (26.15%) | 135 (22.96%) | 148 (25.47%) | 1036 (30.81%) | 1101 (32.41%) |
|  | £30k - £60k | 332 (36.69%) | 362 (40.45%) | 203 (34.52%) | 247 (42.51%) | 1109 (32.99%) | 1165 (34.29%) |
|  | £60k - £90k | 119 (13.15%) | 124 (13.85%) | 105 (17.86%) | 81 (13.94%) | 387 (11.51%) | 410 (12.07%) |
|  | £90k+ | 79 (8.73%) | 93 (10.39%) | 58 (9.86%) | 59 (10.15%) | 215 (6.4%) | 224 (6.59%) |
|  | Openness | 15.1 (3.25) | 15.05 (3.16) | 15.37 (3.17) | 15.08 (3.18) | 15.22 (3.17) | 15.34 (3.17) |
|  | Conscientiousness | 16.36 (2.77) | 16.37 (2.79) | 16.33 (2.79) | 16.33 (2.78) | 16.15 (2.85) | 16.24 (2.89) |
|  | Extraversion | 13.19 (4.15) | 13.22 (4.25) | 13.01 (4.21) | 13.23 (4.3) | 12.86 (4.17) | 13.33 (4.15) |
|  | Agreeableness | 15.82 (2.81) | 15.8 (2.87) | 15.87 (2.94) | 15.78 (2.85) | 15.7 (2.95) | 15.64 (3.01) |
|  | Neuroticism | 10.31 (4.01) | 10.31 (3.86) | 10.44 (4.05) | 10.56 (3.94) | 10.57 (4.12) | 10.36 (4.1) |
| Psychiatric condition | No | 891 (88.92%) | 898 (89.62%) | 559 (86.4%) | 573 (88.56%) | 3306 (87.05%) | 3337 (87.86%) |
|  | Yes | 111 (11.08%) | 104 (10.38%) | 88 (13.6%) | 74 (11.44%) | 492 (12.95%) | 461 (12.14%) |
| Long-Term Conditions | 0 | 505 (50.4%) | 518 (51.7%) | 373 (57.65%) | 342 (52.86%) | 1831 (48.21%) | 1774 (46.71%) |
|  | 1 | 322 (32.14%) | 317 (31.64%) | 184 (28.44%) | 204 (31.53%) | 1223 (32.2%) | 1246 (32.81%) |
|  | 2+ | 175 (17.47%) | 167 (16.67%) | 90 (13.91%) | 101 (15.61%) | 744 (19.59%) | 778 (20.48%) |
| Received Flu Vaccine | No | 233 (23.25%) | 214 (21.36%) | 214 (33.08%) | 179 (27.67%) | 1169 (30.78%) | 1107 (29.15%) |
|  | Yes | 769 (76.75%) | 788 (78.64%) | 433 (66.92%) | 468 (72.33%) | 2629 (69.22%) | 2691 (70.85%) |
|  | Attitude to Vaccines (Factor) | 0.07 (0.93) | 0.1 (0.94) | 0.01 (1.05) | 0.08 (0.97) | 0 (1) | 0.03 (0.94) |
| Vaccination Intention | 1 | 43 (4.29%) | 47 (4.69%) | 46 (7.11%) | 34 (5.26%) | 220 (5.79%) | 180 (4.74%) |
|  | 2 | 30 (2.99%) | 25 (2.5%) | 23 (3.55%) | 18 (2.78%) | 118 (3.11%) | 118 (3.11%) |
|  | 3 | 72 (7.19%) | 62 (6.19%) | 52 (8.04%) | 49 (7.57%) | 267 (7.03%) | 253 (6.66%) |
|  | 4 | 96 (9.58%) | 103 (10.28%) | 73 (11.28%) | 70 (10.82%) | 413 (10.87%) | 404 (10.64%) |
|  | 5 | 169 (16.87%) | 165 (16.47%) | 96 (14.84%) | 108 (16.69%) | 525 (13.82%) | 535 (14.09%) |
|  | 6 | 592 (59.08%) | 600 (59.88%) | 357 (55.18%) | 368 (56.88%) | 2255 (59.37%) | 2308 (60.77%) |
| Shielding | No | 769 (76.75%) | 796 (79.44%) | 544 (84.08%) | 531 (82.07%) | 2923 (76.96%) | 2856 (75.2%) |
|  | Yes | 233 (23.25%) | 206 (20.56%) | 103 (15.92%) | 116 (17.93%) | 875 (23.04%) | 942 (24.8%) |
|  | Propensity Score Distance | 0.3 (0.19) | 0.32 (0.2) | 0.35 (0.27) | 0.37 (0.29) | 0.37 (0.23) | 0.42 (0.25) |
